# Association of Regional Anesthesia with Postoperative Opioid Use After Foot and Ankle Surgery

**DOI:** 10.1101/2025.04.21.25326144

**Authors:** Yuri C. Martins, Joanne Salas, George Tseng, Jeffrey F. Scherrer

**Author notes:** **Corresponding Author:** Yuri C. Martins, MD/PhD, at Department of Anesthesiology, Saint Louis University School of Medicine. Drummond Hall-2nd Floor. 3691 Rutger St. Saint Louis, MO, USA. ZIP 63110. **E-mails:** YCM =; JS =; GT =; JFS =.

## Abstract

**Purpose:** We investigated if the use of peripheral nerve blockade (PNB) was associated with a lower incidence of prescription opioid fill within 30 days post-surgery and persistent postoperative opioid use (PPOU) in patients undergoing foot and ankle surgery.

**Methods:** We identified adults who had undergone foot or ankle surgery between 2012 and 2018 and did or did not receive PNB in an Optum’s de-identified Integrated Claims-Clinical dataset (n=12,643). Pharmacy data was used to track opioid prescription fill date and supply. PPOU was defined as >90 days of continuous opioid use. Entropy balancing was used to control differences in the distribution of covariates. Log-binomial models in unweighted and weighted data estimated crude and adjusted relative risk (RR) with 95% confidence intervals (CI) for the outcomes.

**Results:** One-third of the sample filled an opioid within 30 days of surgery, and among these patients, 57.3% continued use for > 90 days. Performance of PNB was associated with an increased risk for filling opioid prescriptions within 30 days post-surgery before (RR=1.40; 95%CI:1.32-1.49) and after (RR=1.31; 95%CI:1.22-1.41; p<0.0001) controlling for confounding. However, the group that received a PNB showed significantly lower risk of PPOU before (0.91; 95%CI:0.85-0.98; p=0.016) and after controlling for confounding (RR=0.92; 95%CI:0.85-0.99; p=0.029).

**Conclusion:** Performance of PNB for patients undergoing foot and ankle surgery was associated with a 31% increased risk of any opioid prescription fill within 30 days after surgery. However, among the patients that initially filled their prescriptions, patients that received PNB had a significantly (8%) lower risk for PPOU.

## Introduction

The United States of America is amid an opioid crisis that imposes a large burden in terms of morbidity and economic costs [1]. It is estimated that about 40% of opioid overdose deaths involve prescription opioids [2]. Although the past decade has seen steady decreases in opioid prescribing[3], opioid-related adverse events and deaths continue to increase [4, 5]. This increase in opioid-related deaths indicates that new approaches are needed to change the current opioid ecosystem.

Opioid utilization in the perioperative period has been associated with increased risk for persistent pain, addiction, overdose, and respiratory depression [6-10]. In this light, postsurgical opioid prescriptions and the concept of persistent postoperative opioid use (PPOU), typically defined based on filled pharmacy claims for opioids after surgery, emerged as an important focus of opioid-related policy, research, and intervention [10, 11]. Past estimates of the incidence of PPOU use range from 0.01% to 34%, depending on the surgery and population studied [7, 10, 12].

Orthopedic procedures are common and associated with 5-34% risk of PPOU [9, 10, 13-17]. Given concerns related to the relatively high risk of PPOU in orthopedic surgical patients, a question with important clinical implications is whether the use of peripheral nerve blockades (PNBs) can reduce the risk of PPOU. As far as we know, there have been only three studies that have examined if there is an association between PNBs and PPOU in orthopedic surgeries [14-16], which shows the paucity of evidence in this area. Two of these studies focused on ambulatory shoulder surgery [14, 15] and one on total knee arthroplasties [16] with all three studies finding no association between the receipt of a PNB and PPOU. However, this may not be the case for all types of orthopedic surgeries as there are studies showing that nerve blocks can decrease PPOU in thoracotomy and breast cancer surgery [18].

Foot and ankle surgeries are common procedures that can be associated with significant postoperative pain and a relatively high rate of PPOU, at 8.9% [13]. Although most of the time PNBs are used as adjuncts for intra- and post-operative analgesia in shoulder and knee surgeries [19, 20], regional anesthesia can be used as the sole or main anesthetic technique for foot and ankle surgeries [21, 22]. In addition, PNBs have been shown to reduce acute pain, opioid consumption, and opioid related adverse events before discharge in patients undergoing foot and ankle surgery [23, 24]. Therefore, this study aimed to test the hypothesis that the use of PNBs was associated with a lower incidence of prescription opioid fill within 30 days post-surgery and PPOU among patients undergoing foot and ankle surgery. Using administrative claims data, we identified adult patients undergoing foot and ankle surgery between 2012 and 2018. We then determined whether the group of patients who received PNBs had a reduced risk of prescription opioid fill within 30 days post-surgery and PPOU during the first 6-months after surgery.

## Methods

### Subjects

We used a leased Optum’s de-identified Integrated Claims-Clinical data set that contained de-identifed electronic health records (EHR) from a random sample of 5 million adult (≥18 years of age) patients with outpatient or inpatient encounters from the leased database covering 2010 through 2018. The EHR and claims data includes encounters in academic and non-academic healthcare systems throughout the United States and encompasses patients with private, government or no health insurance. Of the 5 million patients obtained from The Integrated data set, approximately 18% were part of Optum’s de-identified Integrated Claims-Clinical dataset (n=897,513), which included data from EHR and medical claims (Figure 1). All study variables were created from data elements in The Integrated data set files. The International Classification of Diseases (ICD)-9-CM and ICD-10-CM codes, procedure codes/claims (ICD-9, -10, and CPT), pharmacy claims, vital signs, laboratory results, demographics and geographic region were used to create study variables. ICD-11 codes were not used. Detailed definitions of all study variables are available in the table in Supplemental Digital Content 1.

**Figure 1.**
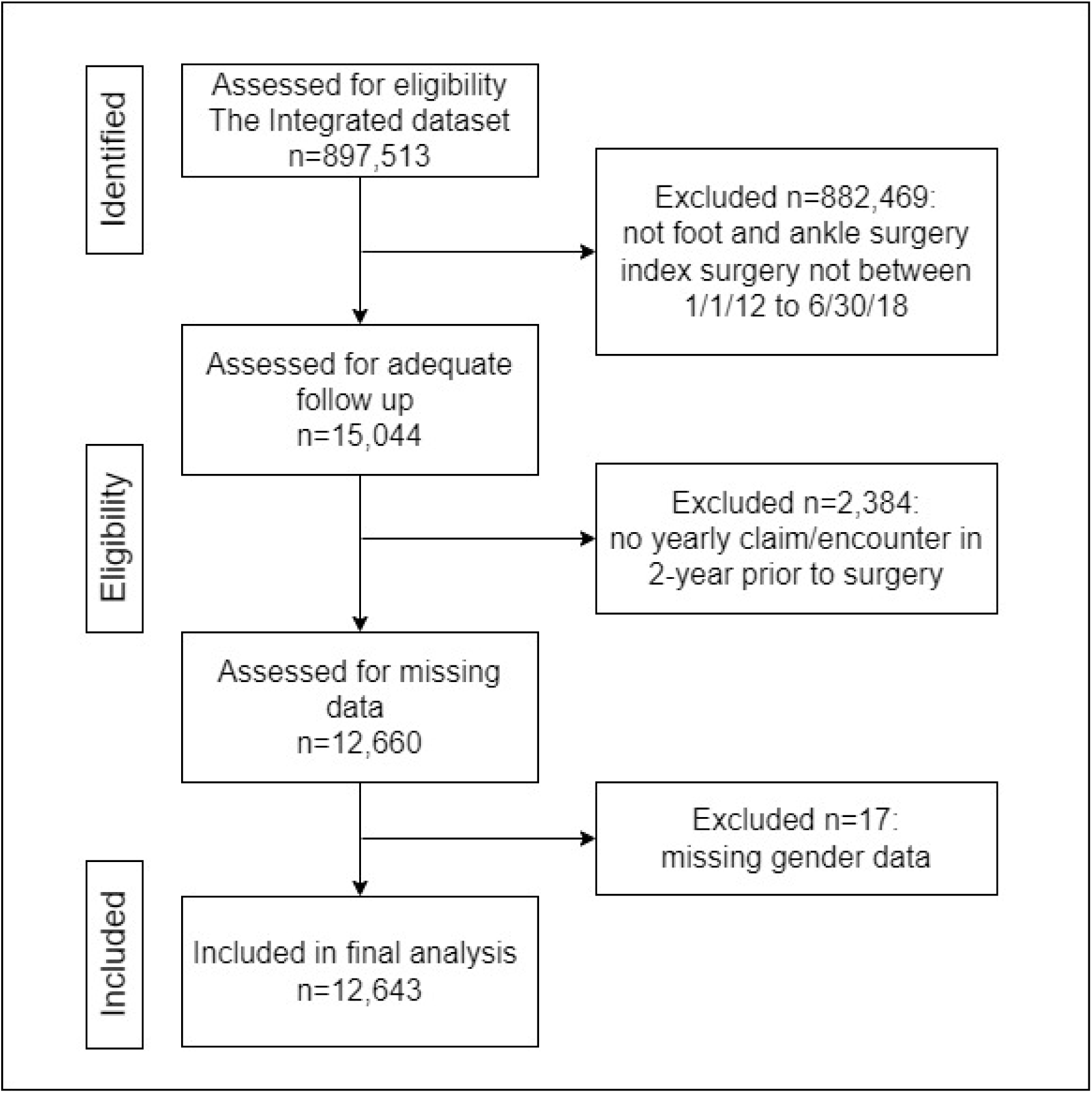
Sampling process from The Integrated Dataset. EHR = electronic health records.

### Eligibility

We selected adult (≥18 years of age) patients who had a claim for foot or ankle surgery between 1/1/2012 and 6/30/2018. The date of surgery was the baseline/index surgery date. This allowed for a two-year lookback to identify the eligible sample and to measure covariates. Also, this allowed for all patients to have a minimum of 6-months of follow-up to measure opioid duration after surgery. To increase the probability that surgery patients would have follow-up care within the same healthcare system, we required an annual encounter or medical claim in the 2-years prior to surgery. After removing 17 patients missing gender data, the analytic sample consisted of 12,643 patients who did or did not receive PNBs during foot or ankle surgery. Figure 1 illustrates the sampling process.

### Outcome

We modeled two separate outcomes. The first was a patient filling any prescription opioid within 30 days post-surgery. The second outcome was PPOU, defined as >90 days of opioid prescription fillings among those who filled an opioid prescription within 30 days of surgery. We considered opioid use to be continuous until a gap of >30 days occurred between fills. Prescription opioids included the following medications in both extended and immediate release formulations: codeine, dihydrocodeine, fentanyl, hydrocodone, hydromorphone, levorphanol, meperidine, methadone, morphine, oxycodone, oxymorphone, pentazocine, tapentadol, and tramadol. While it is possible to determine if methadone was prescribed for opioid use disorder (OUD) or for pain because the former is a procedure and the latter a prescription, it is more difficult to separate buprenorphine prescribed for OUD vs. for pain. Therefore, we do not include buprenorphine in the list of prescription opioids. PPOU was measured using prescription fill date and days’ supply.

### Exposure

The exposure was the application of a peripheral nerve block during foot or ankle surgery. Nerve block was defined by CPT codes 64445, 64446, 64447, 64448, 64450; ICD-9 procedure 04.81; and ICD-10 procedure 3E0T3BZ.

### Covariates

Detailed definitions for all study variables are available in the Supplemental Digital Content 1. Sociodemographic covariates included age, gender, race (White, Black, Other/Unknown), and census region (Northeast, Midwest, South, West, Other/Unknown). Patients who use more healthcare are more likely to receive diagnoses and therapies, we controlled for this detection bias by adjusting for volume of healthcare utilization. We computed the distribution of the average number of visits per month in the 2-years prior to the index surgery and defined high healthcare utilization as the top 25^th^ percentile of the distribution.

All remaining covariates were measured in the 2-years prior to the index surgery, unless otherwise noted. Because past prescription opioid use is associated with future use, we controlled for any opioid fills within the 3-months before surgery. We also controlled for benzodiazepine and gabapentinoid fills because they are frequently co-prescribed with opioids and are associated with long term opioid therapy and receipt of more potent opioids [25]. We adjusted for obesity because it is correlated with chronic pain. Because pre-existing pain conditions may be associated with odds of post-surgical opioid fills, we defined several pain diagnoses from a list of conditions for which an opioid may be prescribed [26]. The pain diagnoses modeled as covariates were arthritis, back/neck pain, chronic pain, fibromyalgia, headache/migraine, muscle pain and neuropathy.

Psychiatric and substance use disorders are associated with increased risk of receiving an opioid and long-term opioid therapy [26]. Therefore, we adjusted for depression and any anxiety disorder, which was defined as anxiety not otherwise specified (NOS), generalized anxiety disorder, panic disorder and/or social phobia. We also controlled for posttraumatic stress disorder, obsessive compulsive disorder, nicotine dependence, alcohol abuse/dependence and any form of drug abuse/dependence.

### Statistical Analysis

Whether a patient receives a PNB for foot or ankle surgery is not random. To control for this bias by indication and for other potential confounding, we used entropy balancing (e-balance). E-balance removes differences in the distribution of covariates between patients with and without a nerve block [27, 28]. E-balance achieves better balance than obtained with propensity scores and inverse probability of treatment weighting because it does not rely on correctly specifying a propensity score model and iteratively checking for balance in the process. E-balance weights the groups such that specified covariate moments (mean, variance) are equal to those with vs. without a PNB. The standardized mean difference percent (SMD%=100*SMD) was used to determine if e-balance was successful. Successful balance is indicated by an SMD% < 10% [29]. The ‘WeightIt’ package in R v4.2.1 was used to calculate e-balance weights.

All remaining analyses were conducted using SAS v9.4 (SAS Institute, Cary, NC). Covariate differences between patients with and without PNBs were assessed using chi-square tests and independent samples t-tests, with SMD% measuring effect size differences before and after e-balance weighting. Log-binomial models in unweighted and weighted data estimated crude and adjusted relative risk (RR) with 95% confidence intervals (CI) of starting an opioid within 30 days of surgery and among those starting an opioid, reaching > 90 days duration. Weighted models used robust, sandwich-type variance estimators to calculate variance and confidence interval [29].

## Results

Baseline cohort characteristics are shown in Table 1. The average age was 56.3 (SD±15.2) years, 67.1% were female, 77.1% were White race and 6.6% were Black. The sample was disproportionately from the South and Midwest. Nearly 1 out of 4 patients filled an opioid prescription in the 3 months prior to surgery. The most common comorbid pain conditions were muscle pain (84.6%) and arthritis (79.9%). Almost one-third of patients were obese. Nicotine dependence was the most prevalent psychiatric disorder (22.6%) followed by depression (17.5%). Less than 4% of the sample had diagnoses for alcohol or drug abuse/dependence.

**Table 1.** Baseline characteristics of ≥ 18 year old patients undergoing foot and ankle surgery.

| Covariates, *N*(%) or mean(±SD) | Total (*N* = 12,643) |
| --- | --- |
| <u>*Sociodemographic-related*</u> |  |
| Age, mean (±sd) | 56.3 (±15.2) |
| Female gender | 8,482 (67.1%) |
| Race |  |
| White | 9,744 (77.1%) |
| Black | 837 (6.6%) |
| Other/Unknown | 2,062 (16.3%) |
| Region |  |
| Midwest | 4,626 (36.6%) |
| Northeast | 2,248 (17.8%) |
| South | 4,111 (32.5%) |
| West | 1,288 (10.2%) |
| Other/unknown | 370 (2.9%) |
| High utilization | 3,160 (25.0%) |
| <u>*Comorbidities*</u> |  |
| Prior opioid fill (3-months before surgery) | 3,062 (24.2%) |
| Arthritis | 10,096 (79.9%) |
| Back/neck pain | 5,852 (46.3%) |
| Chronic pain | 1,144 (9.1%) |
| Fibromyalgia | 1,304 (10.3%) |
| Headache | 2,607 (20.6%) |
| Muscle pain | 10,690 (84.6%) |
| Neuropathy | 3,069 (13.9%) |
| Obesity | 4,158 (32.9%) |
| Benzodiazepine fill | 1,756 (13.9%) |
| Anxiety | 1,986 (15.7%) |
| PTSD | 133 (1.1%) |
| OCD | 52 (0.4%) |
| Depression | 2,212 (17.5%) |
| Nicotine dependence | 2,854 (22.6%) |
| Alcohol abuse/dependence | 415 (3.3%) |
| Drug abuse/dependence | 349 (2.8%) |

*N* = number of patients, SD = standard deviation

The covariate distributions by whether or not patients received a PNB are shown in Table 2. Patients who received PNBs were meaningfully younger than those who did not (SMD% = - 23.6). Those without a PNB, compared to those with a nerve block, were disproportionately from the Northeast (SMD%= -12.8). Opioid fills within the 3-months prior to surgery were more prevalent in patients who had PNBs, compared to those without a nerve block (SMD%=10.6).

**Table 2.** Baseline characteristics of ≥ 18 year old patients, by whether nerve block was received at time of foot and ankle surgery (N=12,643)

| Covariates, N(%) or mean(±SD) | No<br>N = 11,015 | Yes<br>N = 1,628 | P value | SMD (%) |
| --- | --- | --- | --- | --- |
| *<u>Sociodemographic-related</u>* |  |  |  |  |
| Age, mean (±SD) | 56.8 (±15.3) | 53.3 (±14.0) | <.0001 | -23.6 |
| Female gender | 7,444 (67.6%) | 1,038 (63.8%) | .002 | -8.1 |
| Race |  |  |  |  |
| White | 8,474 (76.9%) | 1,270 (78.0%) | .625 | 2.6 |
| Black | 733 (6.7%) | 104 (6.4%) |  | -1.1 |
| Other/Unknown | 1,808 (16.4%) | 254 (15.6%) |  | -2.2 |
| Region |  |  |  |  |
| Midwest | 4,038 (36.7%) | 588 (36.1%) | <.0001 | -1.1 |
| Northeast | 2,025 (18.4%) | 223 (13.7%) |  | -12.8 |
| South | 3,528 (32.0%) | 583 (35.8%) |  | 8.0 |
| West | 1,107 (10.1%) | 181 (11.1%) |  | 3.5 |
| Other/unknown | 317 (2.9%) | 53 (3.3%) |  | 2.2 |
| High utilization | 2,758 (25.0%) | 402 (24.7%) | .764 | -0.8 |
| *<u>Comorbidities</u>* |  |  |  |  |
| Prior opioid fill (3-months before surgery) |  |  |  |  |
|  | 2,602 (23.6%) | 460 (28.3%) | <.0001 | 10.6 |
| Arthritis | 8,606 (78.1%) | 1,490 (91.5%) | <.0001 | 38.0 |
| Back/neck pain | 5,105 (46.4%) | 747 (45.9%) | .727 | -0.9 |
| Chronic pain | 951 (8.6%) | 193 (11.9%) | <.0001 | 10.6 |
| Fibromyalgia | 1,145 (10.4%) | 159 (9.8%) | .436 | -2.1 |
| Headache | 2,264 (20.6%) | 343 (21.1%) | .632 | 1.3 |
| Muscle pain | 9,229 (83.8%) | 1,461 (89.7%) | <.0001 | 17.6 |
| Neuropathy | 2,725 (24.7%) | 344 (21.1%) | .002 | -8.6 |
| Obesity | 3,535 (32.1%) | 623 (38.3%) | <.0001 | 13.0 |
| Benzodiazepine fill | 1,539 (14.0%) | 217 (13.3%) | .484 | -1.9 |
| Anxiety | 1,727 (15.7%) | 259 (15.9%) | .811 | 0.6 |
| PTSD | 108 (1.0%) | 25 (1.5%) | .040 | 5.0 |
| OCD | 42 (0.4%) | 10 (0.6%) | .170 | 3.3 |
| Depression | 1,915 (17.4%) | 297 (18.2%) | .395 | 2.2 |
| Nicotine dependence | 2,485 (22.6%) | 369 (22.7%) | .924 | 0.3 |
| Alcohol abuse/dependence | 375 (3.4%) | 40 (2.5%) | .045 | -5.6 |
| Drug abuse/dependence | 297 (2.7%) | 52 (3.2%) | .252 | 2.9 |

N = number of patients, SD = standard deviation, PTSD = Post-traumatic stress disorder, OCD = Obsessive-compulsive disorder

The diagnosis of arthritis, chronic pain, and muscle pain were each more prevalent in patients that did vs. did not receive a PNB (SMD% range:10.6-38.0). Obesity was more common in patients who had PNBs, compared to those who did not (SMD%=13.0). The distribution of psychiatric and substance use disorders did not differ by nerve block status. After e-balancing, all covariates were balanced between those who did vs. did not receive a PNB (all SMD% < 0.05%).

As shown in Table 3, 44.4% of patients with a PNB, compared to 31.7% of patients without regional anesthesia exposure filled an opioid prescription within 30 days after surgery. Prior to controlling for confounding, PNB was associated with a 40% increased risk for filling any post-operative opioid prescription (RR=1.40; 95%CI:1.32-1.49; p<0.0001). After controlling for confounding by e-balance weighting, this association was slightly attenuated, but PNB remained significantly associated with any post-operative opioid fill (RR=1.31; 95%CI:1.22-1.41; p<0.0001).

**Table 3.** Association between nerve block during foot and ankle surgery and relative risk of any opioid fills within 30 days after surgery. Results from log-binomial models. (*N*=12,643)

| Nerve block status | Any opioid fills, N(%) | Crude RR (95% CI) | Weighted RR (95% CI) |
| --- | --- | --- | --- |
| Overall | 4,217 (33.3%) | --- | --- |
| <u>Nerve block</u> |  |  |  |
| No<br>N = 11,015 | 3,494 (31.7%) | 1.00 | 1.00 |
| Yes<br>N = 1,628 | 723 (44.4%) | 1.40 (1.32 to 1.49)<sup>a</sup> | 1.31 (1.22 to 1.41)<sup>b</sup> |

N = Number of patients; RR = relative risk; CI = confidence interval
<sup>a</sup> Crude P value <0.0001
<sup>b</sup> Weighted P value <0.0001

As shown in Table 4, 19.11% (n=2416) of patients from our initial cohort (n=12643) filled opioid prescriptions continuously for >90 days after their procedures, which meets our definition of PPOU. Among patients that filled an opioid prescription in the first 30 days after surgery (n=4,217), 58.2% of patients that did not receive a PNB, compared to 53.1% of patients with regional anesthesia exposure met our criteria for PPOU. The group that received a PNB was associated with significantly lower risk of PPOU (RR=0.91; 95%CI:0.85-0.98; p=0.016) when compared to those without regional anesthesia (Table 4). This association was nearly unchanged after controlling for confounding (RR=0.92; 95%CI:0.85-0.99; p=0.029) (Table 4).

**Table 4.** Duration of opioid use, <u>if have an opioid fill.</u> Results from log-binomial models. (N = 4,217)

| Nerve block status | Opioid > 90 days, N(%) | Crude RR (95% CI) | Weighted RR (95% CI) |
| --- | --- | --- | --- |
| Overall | 2,416 (57.3%) | --- | --- |
| <u>Nerve block</u> |  |  |  |
| No<br>N = 3,494 | 2,032 (58.2%) | 1.00 | 1.00 |
| Yes<br>N = 723 | 384 (53.1%) | 0.91 (0.85 to 0.98)<sup>a</sup> | 0.92 (0.85 to 0.99)<sup>b</sup> |

N = Number of patients; RR = relative risk; CI = confidence interval
<sup>a</sup> Crude P value = 0.016
<sup>b</sup> Weighted P value = 0.029

## Discussion

In this large, nationally distributed, patient cohort undergoing foot or ankle surgery, receipt of PNBs were associated with a 31% increased risk of any opioid prescription fill within 30 days after surgery. However, among the patients that initially filled their prescriptions, patients that received a PNB had a significantly (8%) lower risk for PPOU when compared to those without exposure to regional anesthesia. Thus, PNB is positively associated with filling an opioid in the initial postoperative period but is inversely associated with PPOU. These associations were independent of pre-operative opioid fills and other numerous risk factors for long term opioid therapy such as depression and substance use disorders.

Our initial finding that PNBs were associated with a 31% increased risk of any opioid prescription fill within 30 days after foot and ankle surgery are in disagreement with a growing body of studies showing a significant decrease in early postoperative opioid consumption and pain scores in patients undergoing foot and ankle surgery that receive regional anesthesia when compared to general/neuraxial anesthesia without a PNB [22]. Most of the studies showing a decrease in opioid use associated with regional anesthesia are single center studies that measured the actual consumption of opioids prescribed [22, 30]. Single center studies capture common practices of a single region or site, differing from the multicenter level approach captured in our dataset. Therefore, our results are important because they are not dependent on the common practices of a single region or hospital. In addition, a recent study evaluating a multi-state administrative claims dataset for opioid prescription fills in anterior cruciate ligament reconstruction surgeries, an approach similar to ours, also showed PNBs being associated with increased odds of opioid prescription fill < 30 days after surgery [31]. Although speculative, our observation suggests that either there is no reduction in postoperative pain after PNBs wore off, or surgeons do not change their prescribing patterns based on whether this intervention is undertaken.

Foot and ankle surgeries can be associated with severe postoperative pain within the first 24 – 48hs after the procedure [13, 22]. The average analgesic duration after single-injection PNBs with ropivacaine or bupivacaine, the most used local anesthetics for these procedures in the USA, is 11 – 18hs [22, 32], which sometimes falls short of providing satisfactory postoperative pain relief even for the first night after the surgery. In addition, after resolution of PNBs, there may be a relatively rapid increase in the severity of pain with some patients reporting excruciating pain and major distress, which is commonly referred to as “rebound pain” [33-35]. Rebound pain can occur in up to 50% of patients that receive PNBs and is independently associated with young age, female sex, body mass index, and bone surgery [34, 35].

Interestingly, in our study, patients that received PNBs were meaningfully younger and had a higher incidence of obesity than patients without a nerve block. In addition, due to the known high incidence of rebound pain in bone surgeries, it is common practice between anesthesiologists to advise orthopedic ambulatory surgery patients to take opioid pain medications as soon as they feel the firsts signs and symptoms that their PNB is wearing off [35]. Therefore, the presence of rebound pain in patients that received PNBs could be one factor associated with their higher incidence of opioid prescription filling in the first 30 days after surgery when compared with patients that did not receive regional anesthesia.

The PPOU incidence of 19.11% in our work is higher than one previous study showing an incidence of 8.9% [13] in foot and ankle surgeries. However, this high PPOU incidence agrees with previous studies showing 5-34% risk of PPOU in orthopedic surgeries [9, 10, 13-17].

Our findings also indicate that PNBs significantly (8%) lower the risk for PPOU in patients undergoing foot and ankle surgery. Our study indicates, on a multicenter level, that the use of PNBs may reduce long-term opioid use in patients undergoing foot and ankle surgery. Although small in intensity (8%), we believe this decrease to be clinically significant due to the high and increasing number of foot and ankle surgeries performed in the USA and the increasing popularity of PNBs in orthopedic surgeries in general [22, 36]. Although rare, PNBs can have complications and increase the total cost of anesthesia [19-21, 35], so their use needs to be also based on multicenter level data as used in the present study.

Three previous studies found no association between PPOU and the receipt of PNBs in patients undergoing ambulatory shoulder surgery [14, 15] and total knee arthroplasty [16]. The past three studies and ours used definitions of persistent postoperative opioid use that differed among each other. Sun and colleagues [16] defined persistent postoperative opioid use as having filled 10 or more prescriptions or >120 days’ supply within the first year of surgery, excluding the first 90 postoperative days (ie, postoperative days 91–365). Mueller and colleagues [15] definition was having filled at least 1 prescription for an opioid between postoperative days 91 and 365.

Hamilton and colleagues [14] had a more complex definition. Specifically, for opioid-naive patients, persistent postoperative opioid prescription fulfillment was classified as a fulfillment of an opioid prescription if an individual fulfilled an opioid prescription of at least a 60-day supply during postoperative days 90 to 365. For preoperative opioid-exposed or -tolerant individuals, persistent postoperative opioid prescription fulfillment is classified as a fulfillment of an opioid prescription if an individual experienced any increase in opioid prescription fulfillment from postoperative days 90 to 365 relative to their baseline use in the 90 days before surgery.

There is no consensus in the literature about the best definition of PPOU. Jivraj and colleagues identified 29 unique definitions in a systematic review of the literature [10]. Their group also showed that the incidence of persistent postoperative opioid use can vary more than 100-fold depending on the definition used when applied to the same cohort of patients [10]. We used a stringent definition that required >90 days of continuous prescribing, shown to have high levels of agreement with other stringent definitions [10], as persistent opioid use often implies and is interpreted as continuous use. We also aimed to increase specificity to avoid overestimating the risk associated with surgery. Therefore, differences in PPOU definition is one factor that could explain why our findings differ from these previous studies.

There were multiple limitations to our study, many of which are inherent to administrative databases research. We were unable to perform detailed chart reviews to verify the accuracy of coding. Because these codes dictate payment, their presence is likely to be specific. However, sensitivity may be low particularly because anesthesia providers cannot bill for a PNB if the block was only intended to provide anesthesia for the procedure itself and was not intended to provide postoperative analgesia. Nevertheless, these intrinsic characteristics would misclassify patients who received PNBs as part of the “no block” group, which would bias our results toward finding no association. Like with any observational study, we also cannot exclude the possibility that residual confounding could have biased our results.

It is also important to recognize that we did not have the ability to quantify the amount of opioid consumed by the patients and the presence of side effects derived from its consumption. There is a large body of research indicating that the number of opioid pills a patient takes home does not necessarily correlate with the patient’s opioid consumption [37-40]. In addition, we are only able to examine whether a PNB was provided, but cannot evaluate whether it worked, its duration, what technique was used, if peripheral nerve catheters were used, or which medications were administered at the time of surgery. We also did not differentiate between emergency versus elective surgeries and we also do not have data specific to surgical and pain outcomes between the different groups, so we cannot comment on the impact of PNBs on other important patient outcomes such as postoperative pain severity, patient functioning and patient satisfaction. Data available to us ended in 2018 and since then post-surgical prescribing practices may have changed across the USA. Thus, results may not generalize to all current post-surgical pain management practices. Finally, as this study used data only from the continental USA, the findings may not generalize to other countries or jurisdictions.

## Conclusion

In summary, we used a large, nationally distributed, Integrated Claims-Clinical dataset to demostrate that performance of PNBs for patients undergoing foot and ankle surgery was associated with a 31% increased risk of any opioid prescription fill within 30 days after surgery. However, unlike previous research, our study found that, on a multicenter level, the use of PNBs is associated with a modest 8% decreased incidence of persistent postoperative opioid use in patients undergoing foot and ankle surgeries.

## Supporting information

Supplemental Digital Content 1

## Supplementary materials

Supplementary table with detailed descriptions of all study variables.

## Data availability

The data that support the findings of this study are available from Optum (www.optum.com). Restrictions apply to the availability of these data, which were used under license for this study.

