## Supplemental Digital Content 1 for "Association of Regional Anesthesia with Postoperative Opioid Use After Foot and Ankle Surgery"

**SUPPLEMENTARY DIGITAL CONTENT - 1**

| **Variable definitions** | |
| --- | --- |
| **Variable** | **Definition** |
| **Foot and ankle surgery** | |
| Foot and ankle surgery | CPT: 01462; 01464; 01470; 01472; 01474; 01480; 01482; 01484; 01486; 01490; 01500; 01520; 01522; 27600; 27601; 27602; 27603; 27604; 27605; 27606; 27607; 27610; 27612; 27613; 27614; 27615; 27616; 27618; 27632; 27619; 27634; 27620; 27625; 27626; 27630; 27632; 27634; 27635; 27637; 27638; 27640; 27641; 27645; 27646; 27647; 27648; 27650; 27652; 27654; 27656; 27658; 27659; 27664; 27665; 27675; 27676; 27680; 27681; 27685; 27686; 27687; 27690; 27691; 27692; 27695; 27696; 27698; 27700; 27702; 27703; 27704; 27705; 27707; 27709; 27712; 27715; 27720; 27722; 27724; 27725; 27726; 27727; 27730; 27732; 27734; 27740; 27742; 27745; 27750; 27752; 27756; 27758; 27759; 27760; 27762; 27766; 27767; 27768; 27769; 27780; 27781; 27784; 27786; 27788; 27792; 27808; 27810; 27814; 27816; 27818; 27822; 27823; 27824; 27825; 27826; 27827; 27828; 27829; 27830; 27831; 27832; 27840; 27842; 27846; 27848; 27860; 27870; 27871; 27880; 27881; 27882; 27884; 27886; 27888; 27889; 27892; 27893; 27894; 27899; 28001; 28002; 28003; 28005; 28008; 28010; 28011; 28020; 28022; 28024; 28035; 28039; 28041; 28043; 28039; 28045; 28041; 28046; 28047; 28050; 28052; 28054; 28055; 28060; 28062; 28070; 28072; 28080; 28086; 28088; 28090; 28092; 28100; 28102; 28103; 28104; 28106; 28107; 28108; 28110; 28111; 28112; 28113; 28114; 28116; 28118; 28119; 28120; 28122; 28124; 28126; 28130; 28140; 28150; 28153; 28160; 28171; 28173; 28175; 28190; 28192; 28193; 28200; 28202; 28208; 28210; 28220; 28222; 28225; 28226; 28230; 28232; 28234; 28238; 28240; 28250; 28260; 28261; 28262; 28264; 28270; 28272; 28280; 28285; 28286; 28288; 28289; 28291; 28292; 28295; 28296; 28295; 28297; 28298; 28299; 28300; 28302; 28304; 28305; 28306; 28307; 28308; 28309; 28310; 28312; 28313; 28315; 28320; 28322; 28340; 28341; 28344; 28345; 28360; 28400; 28405; 28406; 28415; 28420; 28430; 28435; 28436; 28445; 28446; 28450; 28455; 28456; 28465; 28470; 28475; 28476; 28485; 28490; 28495; 28496; 28505; 28510; 28515; 28525; 28530; 28531; 28540; 28545; 28546; 28555; 28570; 28575; 28576; 28585; 28600; 28605; 28606; 28615; 28630; 28635; 28636; 28645; 28660; 28665; 28666; 28675; 28705; 28715; 28725; 28730; 28735; 28737; 28740; 28750; 28755; 28760; 28800; 28805; 28810; 28820; 28825; 28890; 28899;  ICD-9 procedure: 77.07; 77.08; 77.17; 77.18; 77.27; 77.28; 77.37; 77.38; 77.47; 77.48; 77.51; 77.54; 77.56; 77.57; 77.58; 77.59; 77.67; 77.68; 77.77; 77.78; 77.87; 77.88; 77.97; 77.98; 78.07; 78.08; 78.17; 78.18; 78.27; 78.28; 78.37; 78.38; 78.47; 78.48; 78.57; 78.58; 78.67; 78.68; 78.77; 78.78; 78.87; 78.88; 78.97; 78.98; 79.06; 79.07; 79.08; 79.16; 79.17; 79.18; 79.26; 79.27; 79.28; 79.36; 79.37; 79.38; 79.46; 79.56; 79.66; 79.67; 79.68; 79.78; 79.87; 79.88; 79.96; 79.97; 79.98; 80.07; 80.08; 80.17; 80.18; 80.27; 80.28; 80.37; 80.38; 80.47; 80.48; 80.77; 80.78; 80.87; 80.88; 80.97; 80.98; 81.1; 81.11; 81.12; 81.13; 81.14; 81.15; 81.16; 81.17; 81.18; 81.49; 81.56; 81.57; 81.94; 83.11; 83.84; 84.11; 84.12; 84.13; 84.14; 84.15; 84.26; 84.27;  ICD-10 procedure: 0QCG0ZZ; 0QCG3ZZ; 0QCG4ZZ; 0QCH0ZZ; 0QCH3ZZ; 0QCH4ZZ; 0QCJ0ZZ; 0QCJ3ZZ; 0QCJ4ZZ; 0QCK0ZZ; 0QCK3ZZ; 0QCK4ZZ; 0QCL0ZZ; 0QCL3ZZ; 0QCL4ZZ; 0QCM0ZZ; 0QCM3ZZ; 0QCM4ZZ; 0QCN0ZZ; 0QCN3ZZ; 0QCN4ZZ; 0QCP0ZZ; 0QCP3ZZ; 0QCP4ZZ; 0Q9G00Z; 0Q9G0ZZ; 0Q9G40Z; 0Q9G4ZZ; 0Q9H00Z; 0Q9H0ZZ; 0Q9H40Z; 0Q9H4ZZ; 0Q9J00Z; 0Q9J0ZZ; 0Q9J40Z; 0Q9J4ZZ; 0Q9K00Z; 0Q9K0ZZ; 0Q9K40Z; 0Q9K4ZZ; 0Q9L00Z; 0Q9L0ZZ; 0Q9L40Z; 0Q9L4ZZ; 0Q9M00Z; 0Q9M0ZZ; 0Q9M40Z; 0Q9M4ZZ; 0Q9N00Z; 0Q9N0ZZ; 0Q9N40Z; 0Q9N4ZZ; 0Q9P00Z; 0Q9P0ZZ; 0Q9P40Z; 0Q9P4ZZ; 0Q8G0ZZ; 0Q8G3ZZ; 0Q8G4ZZ; 0Q8H0ZZ; 0Q8H3ZZ; 0Q8H4ZZ; 0Q8J0ZZ; 0Q8J3ZZ; 0Q8J4ZZ; 0Q8K0ZZ; 0Q8K3ZZ; 0Q8K4ZZ; 0Q8L0ZZ; 0Q8L3ZZ; 0Q8L4ZZ; 0Q8M0ZZ; 0Q8M3ZZ; 0Q8M4ZZ; 0Q8N0ZZ; 0Q8N3ZZ; 0Q8N4ZZ; 0Q8P0ZZ; 0Q8P3ZZ; 0Q8P4ZZ; 0Q8G0ZZ; 0Q8G3ZZ; 0Q8G4ZZ; 0Q8H0ZZ; 0Q8H3ZZ; 0Q8H4ZZ; 0Q8J0ZZ; 0Q8J3ZZ; 0Q8J4ZZ; 0Q8K0ZZ; 0Q8K3ZZ; 0Q8K4ZZ; 0Q8L0ZZ; 0Q8L3ZZ; 0Q8L4ZZ; 0Q8M0ZZ; 0Q8M3ZZ; 0Q8M4ZZ; 0Q8N0ZZ; 0Q8N3ZZ; 0Q8N4ZZ; 0Q8P0ZZ; 0Q8P3ZZ; 0Q8P4ZZ; 0Q9G0ZX; 0Q9G3ZX; 0Q9G4ZX; 0Q9H0ZX; 0Q9H3ZX; 0Q9H4ZX; 0Q9J0ZX; 0Q9J3ZX; 0Q9J4ZX; 0Q9K0ZX; 0Q9K3ZX; 0Q9K4ZX; 0QBG0ZX; 0QBG3ZX; 0QBG4ZX; 0QBH0ZX; 0QBH3ZX; 0QBH4ZX; 0QBJ0ZX; 0QBJ3ZX; 0QBJ4ZX; 0QBK0ZX; 0QBK3ZX; 0QBK4ZX; 0Q9L0ZX; 0Q9L3ZX; 0Q9L4ZX; 0Q9M0ZX; 0Q9M3ZX; 0Q9M4ZX; 0Q9N0ZX; 0Q9N3ZX; 0Q9N4ZX; 0Q9P0ZX; 0Q9P3ZX; 0Q9P4ZX; 0QBL0ZX; 0QBL3ZX; 0QBL4ZX; 0QBM0ZX; 0QBM3ZX; 0QBM4ZX; 0QBN0ZX; 0QBN3ZX; 0QBN4ZX; 0QBP0ZX; 0QBP3ZX; 0QBP4ZX; 0MQS0ZZ; 0MQS3ZZ; 0MQS4ZZ; 0Q8N0ZZ; 0Q8N3ZZ; 0Q8N4ZZ; 0QBN0ZZ; 0QBN3ZZ; 0QBN4ZZ; 0MQT0ZZ; 0MQT3ZZ; 0MQT4ZZ; 0Q8P0ZZ; 0Q8P3ZZ; 0Q8P4ZZ; 0QBP0ZZ; 0QBP3ZZ; 0QBP4ZZ; 0QBN0ZZ; 0QBN3ZZ; 0QBN4ZZ; 0QBP0ZZ; 0QBP3ZZ; 0QBP4ZZ; 0QBQ0ZZ; 0QBQ3ZZ; 0QBQ4ZZ; 0QBR0ZZ; 0QBR3ZZ; 0QBR4ZZ; 0SGP04Z; 0SGP05Z; 0SGP07Z; 0SGP0JZ; 0SGP0KZ; 0SGP34Z; 0SGP35Z; 0SGP37Z; 0SGP3JZ; 0SGP3KZ; 0SGP44Z; 0SGP45Z; 0SGP47Z; 0SGP4JZ; 0SGP4KZ; 0SGQ04Z; 0SGQ05Z; 0SGQ07Z; 0SGQ0JZ; 0SGQ0KZ; 0SGQ34Z; 0SGQ35Z; 0SGQ37Z; 0SGQ3JZ; 0SGQ3KZ; 0SGQ44Z; 0SGQ45Z; 0SGQ47Z; 0SGQ4JZ; 0SGQ4KZ; 0LQV0ZZ; 0LQV3ZZ; 0LQV4ZZ; 0LQW0ZZ; 0LQW3ZZ; 0LQW4ZZ; 0QBQ0ZZ; 0QBQ3ZZ; 0QBQ4ZZ; 0QBR0ZZ; 0QBR3ZZ; 0QBR4ZZ; 0SGP04Z; 0SGP05Z; 0SGP07Z; 0SGP0JZ; 0SGP0KZ; 0SGP34Z; 0SGP35Z; 0SGP37Z; 0SGP3JZ; 0SGP3KZ; 0SGP44Z; 0SGP45Z; 0SGP47Z; 0SGP4JZ; 0SGP4KZ; 0SGQ04Z; 0SGQ05Z; 0SGQ07Z; 0SGQ0JZ; 0SGQ0KZ; 0SGQ34Z; 0SGQ35Z; 0SGQ37Z; 0SGQ3JZ; 0SGQ3KZ; 0SGQ44Z; 0SGQ45Z; 0SGQ47Z; 0SGQ4JZ; 0SGQ4KZ; 0QQQ0ZZ; 0QQQ3ZZ; 0QQQ4ZZ; 0QQQXZZ; 0QQR0ZZ; 0QQR3ZZ; 0QQR4ZZ; 0QQRXZZ; 0SRP0JZ; 0SRQ0JZ; 0STP0ZZ; 0STQ0ZZ; 0Q5G0ZZ; 0Q5G3ZZ; 0Q5G4ZZ; 0Q5H0ZZ; 0Q5H3ZZ; 0Q5H4ZZ; 0Q5J0ZZ; 0Q5J3ZZ; 0Q5J4ZZ; 0Q5K0ZZ; 0Q5K3ZZ; 0Q5K4ZZ; 0QBG0ZZ; 0QBG3ZZ; 0QBG4ZZ; 0QBH0ZZ; 0QBH3ZZ; 0QBH4ZZ; 0QBJ0ZZ; 0QBJ3ZZ; 0QBJ4ZZ; 0QBK0ZZ; 0QBK3ZZ; 0QBK4ZZ; 0Q5L0ZZ; 0Q5L3ZZ; 0Q5L4ZZ; 0Q5M0ZZ; 0Q5M3ZZ; 0Q5M4ZZ; 0Q5N0ZZ; 0Q5N3ZZ; 0Q5N4ZZ; 0Q5P0ZZ; 0Q5P3ZZ; 0Q5P4ZZ; 0QBL0ZZ; 0QBL3ZZ; 0QBL4ZZ; 0QBM0ZZ; 0QBM3ZZ; 0QBM4ZZ; 0QBN0ZZ; 0QBN3ZZ; 0QBN4ZZ; 0QBP0ZZ; 0QBP3ZZ; 0QBP4ZZ; 0QBG0ZZ; 0QBG3ZZ; 0QBG4ZZ; 0QBH0ZZ; 0QBH3ZZ; 0QBH4ZZ; 0QBJ0ZZ; 0QBJ3ZZ; 0QBJ4ZZ; 0QBK0ZZ; 0QBK3ZZ; 0QBK4ZZ; 0QBL0ZZ; 0QBL3ZZ; 0QBL4ZZ; 0QBM0ZZ; 0QBM3ZZ; 0QBM4ZZ; 0QBN0ZZ; 0QBN3ZZ; 0QBN4ZZ; 0QBP0ZZ; 0QBP3ZZ; 0QBP4ZZ; 0QBG0ZZ; 0QBG3ZZ; 0QBG4ZZ; 0QBH0ZZ; 0QBH3ZZ; 0QBH4ZZ; 0QBJ0ZZ; 0QBJ3ZZ; 0QBJ4ZZ; 0QBK0ZZ; 0QBK3ZZ; 0QBK4ZZ; 0QBL0ZZ; 0QBL3ZZ; 0QBL4ZZ; 0QBM0ZZ; 0QBM3ZZ; 0QBM4ZZ; 0QBN0ZZ; 0QBN3ZZ; 0QBN4ZZ; 0QBP0ZZ; 0QBP3ZZ; 0QBP4ZZ; 0QTG0ZZ; 0QTH0ZZ; 0QTJ0ZZ; 0QTK0ZZ; 0QTL0ZZ; 0QTM0ZZ; 0QTN0ZZ; 0QTP0ZZ; 0QRG07Z; 0QRG0KZ; 0QRG37Z; 0QRG3KZ; 0QRG47Z; 0QRG4KZ; 0QRH07Z; 0QRH0KZ; 0QRH37Z; 0QRH3KZ; 0QRH47Z; 0QRH4KZ; 0QRJ07Z; 0QRJ0KZ; 0QRJ37Z; 0QRJ3KZ; 0QRJ47Z; 0QRJ4KZ; 0QRK07Z; 0QRK0KZ; 0QRK37Z; 0QRK3KZ; 0QRK47Z; 0QRK4KZ; 0QUG07Z; 0QUG0KZ; 0QUG37Z; 0QUG3KZ; 0QUG47Z; 0QUG4KZ; 0QUH07Z; 0QUH0KZ; 0QUH37Z; 0QUH3KZ; 0QUH47Z; 0QUH4KZ; 0QUJ07Z; 0QUJ0KZ; 0QUJ37Z; 0QUJ3KZ; 0QUJ47Z; 0QUJ4KZ; 0QUK07Z; 0QUK0KZ; 0QUK37Z; 0QUK3KZ; 0QUK47Z; 0QUK4KZ; 0QRL07Z; 0QRL0KZ; 0QRL37Z; 0QRL3KZ; 0QRL47Z; 0QRL4KZ; 0QRM07Z; 0QRM0KZ; 0QRM37Z; 0QRM3KZ; 0QRM47Z; 0QRM4KZ; 0QRN07Z; 0QRN0KZ; 0QRN37Z; 0QRN3KZ; 0QRN47Z; 0QRN4KZ; 0QRP07Z; 0QRP0KZ; 0QRP37Z; 0QRP3KZ; 0QRP47Z; 0QRP4KZ; 0QUL07Z; 0QUL0KZ; 0QUL37Z; 0QUL3KZ; 0QUL47Z; 0QUL4KZ; 0QUM07Z; 0QUM0KZ; 0QUM37Z; 0QUM3KZ; 0QUM47Z; 0QUM4KZ; 0QUN07Z; 0QUN0KZ; 0QUN37Z; 0QUN3KZ; 0QUN47Z; 0QUN4KZ; 0QUP07Z; 0QUP0KZ; 0QUP37Z; 0QUP3KZ; 0QUP47Z; 0QUP4KZ; 0QHG05Z; 0QHG35Z; 0QHG45Z; 0QHH05Z; 0QHH35Z; 0QHH45Z; 0QHJ05Z; 0QHJ35Z; 0QHJ45Z; 0QHK05Z; 0QHK35Z; 0QHK45Z; 0QHL05Z; 0QHL35Z; 0QHL45Z; 0QHM05Z; 0QHM35Z; 0QHM45Z; 0QHN05Z; 0QHN35Z; 0QHN45Z; 0QHP05Z; 0QHP35Z; 0QHP45Z; 0QBG0ZZ; 0QBG3ZZ; 0QBG4ZZ; 0QBH0ZZ; 0QBH3ZZ; 0QBH4ZZ; 0QBJ0ZZ; 0QBJ3ZZ; 0QBJ4ZZ; 0QBK0ZZ; 0QBK3ZZ; 0QBK4ZZ; 0QBL0ZZ; 0QBL3ZZ; 0QBL4ZZ; 0QBM0ZZ; 0QBM3ZZ; 0QBM4ZZ; 0QBN0ZZ; 0QBN3ZZ; 0QBN4ZZ; 0QBP0ZZ; 0QBP3ZZ; 0QBP4ZZ; 0Q8G0ZZ; 0Q8G3ZZ; 0Q8G4ZZ; 0QRG07Z; 0QRG0KZ; 0QRG37Z; 0QRG3KZ; 0QRG47Z; 0QRG4KZ; 0QUG07Z; 0QUG0KZ; 0QUG37Z; 0QUG3KZ; 0QUG47Z; 0QUG4KZ; 0Q8H0ZZ; 0Q8H3ZZ; 0Q8H4ZZ; 0QRH07Z; 0QRH0KZ; 0QRH37Z; 0QRH3KZ; 0QRH47Z; 0QRH4KZ; 0QUH07Z; 0QUH0KZ; 0QUH37Z; 0QUH3KZ; 0QUH47Z; 0QUH4KZ; 0Q8J0ZZ; 0Q8J3ZZ; 0Q8J4ZZ; 0QRJ07Z; 0QRJ0KZ; 0QRJ37Z; 0QRJ3KZ; 0QRJ47Z; 0QRJ4KZ; 0QUJ07Z; 0QUJ0KZ; 0QUJ37Z; 0QUJ3KZ; 0QUJ47Z; 0QUJ4KZ; 0Q8K0ZZ; 0Q8K3ZZ; 0Q8K4ZZ; 0QRK07Z; 0QRK0KZ; 0QRK37Z; 0QRK3KZ; 0QRK47Z; 0QRK4KZ; 0QUK07Z; 0QUK0KZ; 0QUK37Z; 0QUK3KZ; 0QUK47Z; 0QUK4KZ; 0Q8L0ZZ; 0Q8L3ZZ; 0Q8L4ZZ; 0QRL07Z; 0QRL0KZ; 0QRL37Z; 0QRL3KZ; 0QRL47Z; 0QRL4KZ; 0QUL07Z; 0QUL0KZ; 0QUL37Z; 0QUL3KZ ; 0QUL47Z ; 0QUL4KZ ; 0Q8M0ZZ ;0Q8M3ZZ ;0Q8M4ZZ ;0QRM07Z ;0QRM0KZ ; 0QRM37Z ;0QRM3KZ ; 0QRM47Z ; 0QRM4KZ ; 0QUM07Z ; 0QUM0KZ ; 0QUM37Z ; 0QUM3KZ ; 0QUM47Z ; 0QUM4KZ ; 0Q8N0ZZ ; 0Q8N3ZZ ; 0Q8N4ZZ ; 0QRN07Z ; 0QRN0KZ ; 0QRN37Z ; 0QRN3KZ ; 0QRN47Z ;0QRN4KZ ; 0QUN07Z ; 0QUN0KZ ; 0QUN37Z ; 0QUN3KZ ; 0QUN47Z ;0QUN4KZ ;0Q8P0ZZ ; 0Q8P3ZZ ; 0Q8P4ZZ ; 0QRP07Z ; 0QRP0KZ ; 0QRP37Z ; 0QRP3KZ ; 0QRP47Z ; 0QRP4KZ ; 0QUP07Z ; 0QUP0KZ ; 0QUP37Z ; 0QUP3KZ ; 0QUP47Z ; 0QUP4KZ ; 0QNG0ZZ ; 0QNG3ZZ ; 0QNG4ZZ ; 0QNH0ZZ ; 0QNH3ZZ ; 0QNH4ZZ ; 0QNJ0ZZ ; 0QNJ3ZZ ; 0QNJ4ZZ ; 0QNK0ZZ ; 0QNK3ZZ ; 0QNK4ZZ ; 0QQG0ZZ ; 0QQG3ZZ ; 0QQG4ZZ ; 0QQGXZZ ; 0QQH0ZZ ; 0QQH3ZZ ; 0QQH4ZZ ; 0QQHXZZ ; 0QQJ0ZZ ; 0QQJ3ZZ ; 0QQJ4ZZ ; 0QQJXZZ ; 0QQK0ZZ ; 0QQK3ZZ ; 0QQK4ZZ ; 0QQKXZZ ; 0QUG0JZ ; 0QUG3JZ ; 0QUG4JZ ; 0QUH0JZ ; 0QUH3JZ ; 0QUH4JZ ; 0QUJ0JZ ; 0QUJ3JZ ; 0QUJ4JZ ; 0QUK0JZ ; 0QUK3JZ ; 0QUK4JZ ; 0QNL0ZZ ; 0QNL3ZZ ; 0QNL4ZZ ; 0QNM0ZZ ; 0QNM3ZZ ; 0QNM4ZZ ; 0QNN0ZZ ; 0QNN3ZZ ; 0QNN4ZZ ; 0QNP0ZZ ; 0QNP3ZZ ; 0QNP4ZZ ; 0QQL0ZZ ; 0QQL3ZZ ; 0QQL4ZZ ; 0QQLXZZ ; 0QQM0ZZ ; 0QQM3ZZ ; 0QQM4ZZ ; 0QQMXZZ ; 0QQN0ZZ ; 0QQN3ZZ ; 0QQN4ZZ ; 0QQNXZZ ; 0QQP0ZZ ; 0QQP3ZZ ; 0QQP4ZZ ; 0QQPXZZ ; 0QUL0JZ ; 0QUL3JZ ; 0QUL4JZ ; 0QUM0JZ ; 0QUM3JZ ; 0QUM4JZ ; 0QUN0JZ ; 0QUN3JZ ; 0QUN4JZ ; 0QUP0JZ ; 0QUP3JZ ; 0QUP4JZ ; 0QHG04Z ; 0QHG34Z ; 0QHG44Z ; 0QHH04Z ; 0QHH34Z ; 0QHH44Z ; 0QHJ04Z ; 0QHJ34Z ; 0QHJ44Z ; 0QHK04Z ; 0QHK34Z ; 0QHK44Z ; 0QHL04Z ; 0QHL34Z ; 0QHL44Z ; 0QHM04Z ; 0QHM34Z ; 0QHM44Z ; 0QHN04Z ; 0QHN34Z ; 0QHN44Z ; 0QHP04Z ; 0QHP34Z ; 0QHP44Z ; 0QPG04Z ; 0QPG05Z ; 0QPG34Z ; 0QPG35Z ; 0QPG44Z ; 0QPG45Z ; 0QPH04Z ; 0QPH05Z ; 0QPH34Z ; 0QPH35Z ; 0QPH44Z ; 0QPH45Z ; 0QPJ04Z ; 0QPJ05Z ; 0QPJ34Z ; 0QPJ35Z ; 0QPJ44Z ; 0QPJ45Z ; 0QPK04Z ; 0QPK05Z ; 0QPK34Z ; 0QPK35Z ; 0QPK44Z ; 0QPK45Z ; 0QPL04Z ; 0QPL05Z ; 0QPL34Z ; 0QPL35Z ; 0QPL44Z ; 0QPL45Z ; 0QPM04Z ; 0QPM05Z ; 0QPM34Z ; 0QPM35Z ; 0QPM44Z ; 0QPM45Z ; 0QPN04Z ; 0QPN05Z ; 0QPN34Z ; 0QPN35Z ; 0QPN44Z ; 0QPN45Z ; 0QPP04Z ; 0QPP05Z ; 0QPP34Z ; 0QPP35Z ; 0QPP44Z ; 0QPP45Z ; 0Q8G0ZZ ; 0Q8G3ZZ ; 0Q8G4ZZ ; 0Q8H0ZZ ; 0Q8H3ZZ ; 0Q8H4ZZ ; 0Q8J0ZZ ; 0Q8J3ZZ ; 0Q8J4ZZ ; 0Q8K0ZZ ; 0Q8K3ZZ ; 0Q8K4ZZ ; 0Q8L0ZZ ; 0Q8L3ZZ ; 0Q8L4ZZ ; 0Q8M0ZZ ; 0Q8M3ZZ ; 0Q8M4ZZ ; 0Q8N0ZZ ; 0Q8N3ZZ ; 0Q8N4ZZ ; 0Q8P0ZZ ; 0Q8P3ZZ ; 0Q8P4ZZ ; 0QJY0ZZ ; 0QJY4ZZ ; 0QJY0ZZ ; 0QJY4ZZ ; 0QHY0MZ ; 0QHY3MZ ; 0QHY4MZ ; 0QHY0MZ ; 0QHY3MZ ; 0QHY4MZ ; 0QSD3ZZ ; 0QSD4ZZ ; 0QSDXZZ ; 0QSF3ZZ ; 0QSF4ZZ ; 0QSFXZZ ; 0QSG3ZZ ; 0QSG4ZZ ; 0QSGXZZ ; 0QSH3ZZ ; 0QSH4ZZ ; 0QSHXZZ ; 0QSJ3ZZ ; 0QSJ4ZZ ; 0QSJXZZ ; 0QSK3ZZ ; 0QSK4ZZ ; 0QSKXZZ ; 0QSL3ZZ ; 0QSL4ZZ ; 0QSLXZZ ; 0QSM3ZZ ; 0QSM4ZZ ; 0QSMXZZ ; 0QSN3Z2 ; 0QSN3ZZ ; 0QSN4Z2 ; 0QSN4ZZ ; 0QSNXZ2 ; 0QSNXZZ ; 0QSP3Z2 ; 0QSP3ZZ ; 0QSP4Z2 ; 0QSP4ZZ ; 0QSPXZ2 ; 0QSPXZZ ; 0QSQ3ZZ ; 0QSQ4ZZ ; 0QSQXZZ ; 0QSR3ZZ ; 0QSR4ZZ ; 0QSRXZZ ; 0QSG34Z ; 0QSG44Z ; 0QSH34Z ; 0QSH44Z ; 0QSJ34Z ; 0QSJ44Z ; 0QSK34Z ; 0QSK44Z ; 0QSL34Z ; 0QSL44Z ; 0QSM34Z ; 0QSM44Z ; 0QSN342 ; 0QSN34Z ; 0QSN442 ; 0QSN44Z ; 0QSP342 ; 0QSP34Z ; 0QSP442 ; 0QSP44Z ; 0QSQ34Z ; 0QSQ44Z ; 0QSR34Z ; 0QSR44Z ; 0QSG0ZZ ; 0QSH0ZZ ; 0QSJ0ZZ ; 0QSK0ZZ ; 0QSL0ZZ ; 0QSM0ZZ ; 0QSN0Z2 ; 0QSN0ZZ ; 0QSP0Z2 ; 0QSP0ZZ ; 0QSQ0ZZ ; 0QSR0ZZ ; 0QSG04Z ; 0QSH04Z ; 0QSJ04Z ; 0QSK04Z ; 0QSL04Z ; 0QSM04Z ; 0QSN042 ; 0QSN04Z ; 0QSP042 ; 0QSP04Z ; 0QSQ04Z ; 0QSR04Z ; 0QSG34Z ; 0QSG44Z ; 0QSGXZZ ; 0QSH34Z ; 0QSH44Z ; 0QSHXZZ ; 0QSJ34Z ; 0QSJ44Z ; 0QSJXZZ ; 0QSK34Z ; 0QSK44Z ; 0QSKXZZ ; 0QSG04Z ; 0QSG0ZZ ; 0QSH04Z ; 0QSH0ZZ ; 0QSJ04Z ; 0QSJ0ZZ ; 0QSK04Z ; 0QSK0ZZ ; 0QBG0ZZ ; 0QBH0ZZ ; 0QBJ0ZZ ; 0QBK0ZZ ; 0QBL0ZZ ; 0QBM0ZZ ; 0QBN0ZZ ; 0QBP0ZZ ; 0QBQ0ZZ ; 0QBR0ZZ ; 0SSH34Z ; 0SSH35Z ; 0SSH3ZZ ; 0SSH44Z ; 0SSH45Z ; 0SSH4ZZ ; 0SSHX4Z ; 0SSHX5Z ; 0SSHXZZ ; 0SSJ34Z ; 0SSJ35Z ; 0SSJ3ZZ ; 0SSJ44Z ; 0SSJ45Z ; 0SSJ4ZZ ; 0SSJX4Z ; 0SSJX5Z ; 0SSJXZZ ; 0SSK34Z ; 0SSK35Z ; 0SSK3ZZ ; 0SSK44Z ; 0SSK45Z ; 0SSK4ZZ ; 0SSKX4Z ; 0SSKX5Z ; 0SSKXZZ ; 0SSL34Z ; 0SSL35Z ; 0SSL3ZZ ; 0SSL44Z ; 0SSL45Z ; 0SSL4ZZ ; 0SSLX4Z ; 0SSLX5Z ; 0SSLXZZ ; 0SSM34Z ; 0SSM35Z ; 0SSM3ZZ ; 0SSM44Z ; 0SSM45Z ; 0SSM4ZZ ; 0SSMX4Z ; 0SSMX5Z ; 0SSMXZZ ; 0SSN34Z ; 0SSN35Z ; 0SSN3ZZ ; 0SSN44Z ; 0SSN45Z ; 0SSN4ZZ ; 0SSNX4Z ; 0SSNX5Z ; 0SSNXZZ ; 0SSP34Z ; 0SSP35Z ; 0SSP3ZZ ; 0SSP44Z ; 0SSP45Z ; 0SSP4ZZ ; 0SSPX4Z ; 0SSPX5Z ; 0SSPXZZ ; 0SSQ34Z ; 0SSQ35Z ; 0SSQ3ZZ ; 0SSQ44Z ; 0SSQ45Z ; 0SSQ4ZZ ; 0SSQX4Z ; 0SSQX5Z ; 0SSQXZZ ; 0SSF04Z ; 0SSF05Z ; 0SSF0ZZ ; 0SSG04Z ; 0SSG05Z ; 0SSG0ZZ ; 0SSH04Z ; 0SSH05Z ; 0SSH0ZZ ; 0SSJ04Z ; 0SSJ05Z ; 0SSJ0ZZ ; 0SSK04Z ; 0SSK05Z ; 0SSK0ZZ ; 0SSL04Z ; 0SSL05Z ; 0SSL0ZZ ; 0SSM04Z ; 0SSM05Z ; 0SSM0ZZ ; 0SSN04Z ; 0SSN05Z ; 0SSN0ZZ ; 0SSP04Z ; 0SSP05Z ; 0SSP0ZZ ; 0SSQ04Z ; 0SSQ05Z ; 0SSQ0ZZ ; 0QQG0ZZ ; 0QQG3ZZ ; 0QQG4ZZ ; 0QQGXZZ ; 0QQH0ZZ ; 0QQH3ZZ ; 0QQH4ZZ ; 0QQHXZZ ; 0QQJ0ZZ ; 0QQJ3ZZ ; 0QQJ4ZZ ; 0QQJXZZ ; 0QQK0ZZ ; 0QQK3ZZ ; 0QQK4ZZ ; 0QQKXZZ ; 0QQL0ZZ ; 0QQL3ZZ ; 0QQL4ZZ ; 0QQLXZZ ; 0QQM0ZZ ; 0QQM3ZZ ; 0QQM4ZZ ; 0QQMXZZ ; 0QQN0ZZ ; 0QQN3ZZ ; 0QQN4ZZ ; 0QQNXZZ ; 0QQP0ZZ ; 0QQP3ZZ ; 0QQP4ZZ ; 0QQPXZZ ; 0QQQ0ZZ ; 0QQQ3ZZ ; 0QQQ4ZZ ; 0QQQXZZ ; 0QQR0ZZ ; 0QQR3ZZ ; 0QQR4ZZ ; 0QQRXZZ ; 0SPF0JZ ; 0SPF3JZ ; 0SPF4JZ ; 0SPG0JZ ; 0SPG3JZ ; 0SPG4JZ ; 0SPH0JZ ; 0SPH3JZ ; 0SPH4JZ ; 0SPJ0JZ ; 0SPJ3JZ ; 0SPJ4JZ ; 0SPK0JZ ; 0SPK3JZ ; 0SPK4JZ ; 0SPL0JZ ; 0SPL3JZ ; 0SPL4JZ ; 0SPM0JZ ; 0SPM3JZ ; 0SPM4JZ ; 0SPN0JZ ; 0SPN3JZ ; 0SPN4JZ ; 0SPP0JZ ; 0SPP3JZ ; 0SPP4JZ ; 0SPQ0JZ ; 0SPQ3JZ ; 0SPQ4JZ ; 0M9Q00Z ; 0M9Q0ZZ ; 0M9Q40Z ; 0M9R00Z ; 0M9R0ZZ ; 0M9R40Z ; 0MCQ0ZZ ; 0MCQ3ZZ ; 0MCQ4ZZ ; 0MCR0ZZ ; 0MCR3ZZ ; 0MCR4ZZ ; 0S9F00Z ; 0S9F0ZZ ; 0S9G00Z ; 0S9G0ZZ ; 0SCF0ZZ ; 0SCF3ZZ ; 0SCF4ZZ ; 0SCG0ZZ ; 0SCG3ZZ ; 0SCG4ZZ ; 0SJF0ZZ ; 0SJG0ZZ ; 0M9S00Z ; 0M9S0ZZ ; 0M9S40Z ; 0M9T00Z ; 0M9T0ZZ ; 0M9T40Z ; 0MCS0ZZ ; 0MCS3ZZ ; 0MCS4ZZ ; 0MCT0ZZ ; 0MCT3ZZ ; 0MCT4ZZ ; 0S9H00Z ; 0S9H0ZZ ; 0S9J00Z ; 0S9J0ZZ ; 0S9K00Z ; 0S9K0ZZ ; 0S9L00Z ; 0S9L0ZZ ; 0S9M00Z ; 0S9M0ZZ ; 0S9N00Z ; 0S9N0ZZ ; 0S9P00Z ; 0S9P0ZZ ; 0S9Q00Z ; 0S9Q0ZZ ; 0SCH0ZZ ; 0SCH3ZZ ; 0SCH4ZZ ; 0SCJ0ZZ ; 0SCJ3ZZ ; 0SCJ4ZZ ; 0SCK0ZZ ; 0SCK3ZZ ; 0SCK4ZZ ; 0SCL0ZZ ; 0SCL3ZZ ; 0SCL4ZZ ; 0SCM0ZZ ; 0SCM3ZZ ; 0SCM4ZZ ; 0SCN0ZZ ; 0SCN3ZZ ; 0SCN4ZZ ; 0SCP0ZZ ; 0SCP3ZZ ; 0SCP4ZZ ; 0SCQ0ZZ ; 0SCQ3ZZ ; 0SCQ4ZZ ; 0SJH0ZZ ; 0SJJ0ZZ ; 0SJK0ZZ ; 0SJL0ZZ ; 0SJM0ZZ ; 0SJN0ZZ ; 0SJP0ZZ ; 0SJQ0ZZ ; 0SJF4ZZ ; 0SJG4ZZ ; 0SJH4ZZ ; 0SJJ4ZZ ; 0SJK4ZZ ; 0SJL4ZZ ; 0SJM4ZZ ; 0SJN4ZZ ; 0SJP4ZZ ; 0SJQ4ZZ ; 0M9Q0ZX ; 0M9Q3ZX ; 0M9Q4ZX ; 0M9R0ZX ; 0M9R3ZX ; 0M9R4ZX ; 0MBQ0ZX ; 0MBQ3ZX ; 0MBQ4ZX ; 0MBR0ZX ; 0MBR3ZX ; 0MBR4ZX ; 0S9F0ZX ; 0S9F3ZX ; 0S9F4ZX ; 0S9G0ZX ; 0S9G3ZX ; 0S9G4ZX ; 0SBF0ZX ; 0SBF3ZX ; 0SBF4ZX ; 0SBG0ZX ; 0SBG3ZX ; 0SBG4ZX ; 0M9S0ZX ; 0M9S3ZX ; 0M9S4ZX ; 0M9T0ZX ; 0M9T3ZX ; 0M9T4ZX ; 0MBS0ZX ; 0MBS3ZX ; 0MBS4ZX ; 0MBT0ZX ; 0MBT3ZX ; 0MBT4ZX ; 0S9H0ZX ; 0S9H3ZX ; 0S9H4ZX ; 0S9J0ZX ; 0S9J3ZX ; 0S9J4ZX ; 0S9K0ZX ; 0S9K3ZX ; 0S9K4ZX ; 0S9L0ZX ; 0S9L3ZX ; 0S9L4ZX ; 0S9M0ZX ; 0S9M3ZX ; 0S9M4ZX ; 0S9N0ZX ; 0S9N3ZX ; 0S9N4ZX ; 0S9P0ZX ; 0S9P3ZX ; 0S9P4ZX ; 0S9Q0ZX ; 0S9Q3ZX ; 0S9Q4ZX ; 0SBH0ZX ; 0SBH3ZX ; 0SBH4ZX ; 0SBJ0ZX ; 0SBJ3ZX ; 0SBJ4ZX ; 0SBK0ZX ; 0SBK3ZX ; 0SBK4ZX ; 0SBL0ZX ; 0SBL3ZX ; 0SBL4ZX ; 0SBM0ZX ; 0SBM3ZX ; 0SBM4ZX ; 0SBN0ZX ; 0SBN3ZX ; 0SBN4ZX ; 0SBP0ZX ; 0SBP3ZX ; 0SBP4ZX ; 0SBQ0ZX ; 0SBQ3ZX ; 0SBQ4ZX ; 0M8Q0ZZ ; 0M8Q3ZZ ; 0M8Q4ZZ ; 0M8R0ZZ ; 0M8R3ZZ ; 0M8R4ZZ ; 0MNQ0ZZ ; 0MNQ3ZZ ; 0MNQ4ZZ ; 0MNQXZZ ; 0MNR0ZZ ; 0MNR3ZZ ; 0MNR4ZZ ; 0MNRXZZ ; 0SNF0ZZ ; 0SNF3ZZ ; 0SNF4ZZ ; 0SNG0ZZ ; 0SNG3ZZ ; 0SNG4ZZ ; 0M8S0ZZ ; 0M8S3ZZ ; 0M8S4ZZ ; 0M8T0ZZ ; 0M8T3ZZ ; 0M8T4ZZ ; 0MNS0ZZ ; 0MNS3ZZ ; 0MNS4ZZ ; 0MNSXZZ ; 0MNT0ZZ ; 0MNT3ZZ ; 0MNT4ZZ ; 0MNTXZZ ; 0SNH0ZZ ; 0SNH3ZZ ; 0SNH4ZZ ; 0SNJ0ZZ ; 0SNJ3ZZ ; 0SNJ4ZZ ; 0SNK0ZZ ; 0SNK3ZZ ; 0SNK4ZZ ; 0SNL0ZZ ; 0SNL3ZZ ; 0SNL4ZZ ; 0SNM0ZZ ; 0SNM3ZZ ; 0SNM4ZZ ; 0SNN0ZZ ; 0SNN3ZZ ; 0SNN4ZZ ; 0SNP0ZZ ; 0SNP3ZZ ; 0SNP4ZZ ; 0SNQ0ZZ ; 0SNQ3ZZ ; 0SNQ4ZZ ; 0SBF0ZZ ; 0SBF3ZZ ; 0SBF4ZZ ; 0SBG0ZZ ; 0SBG3ZZ ; 0SBG4ZZ ; 0SBH0ZZ ; 0SBH3ZZ ; 0SBH4ZZ ; 0SBJ0ZZ ; 0SBJ3ZZ ; 0SBJ4ZZ ; 0SBK0ZZ ; 0SBK3ZZ ; 0SBK4ZZ ; 0SBL0ZZ ; 0SBL3ZZ ; 0SBL4ZZ ; 0SBM0ZZ ; 0SBM3ZZ ; 0SBM4ZZ ; 0SBN0ZZ ; 0SBN3ZZ ; 0SBN4ZZ ; 0SBP0ZZ ; 0SBP3ZZ ; 0SBP4ZZ ; 0SBQ0ZZ ; 0SBQ3ZZ ; 0SBQ4ZZ ; 0M5Q0ZZ ; 0M5Q3ZZ ; 0M5Q4ZZ ; 0M5R0ZZ ; 0M5R3ZZ ; 0M5R4ZZ ; 0MBQ0ZZ ; 0MBQ3ZZ ; 0MBQ4ZZ ; 0MBR0ZZ ; 0MBR3ZZ ; 0MBR4ZZ ; 0S5F0ZZ ; 0S5F3ZZ ; 0S5F4ZZ ; 0S5G0ZZ ; 0S5G3ZZ ; 0S5G4ZZ ; 0SBF0ZZ ; 0SBF3ZZ ; 0SBF4ZZ ; 0SBG0ZZ ; 0SBG3ZZ ; 0SBG4ZZ ; 0M5S0ZZ ; 0M5S3ZZ ; 0M5S4ZZ ; 0M5T0ZZ ; 0M5T3ZZ ; 0M5T4ZZ ; 0MBS0ZZ ; 0MBS3ZZ ; 0MBS4ZZ ; 0MBT0ZZ ; 0MBT3ZZ ; 0MBT4ZZ ; 0S5H0ZZ ; 0S5H3ZZ ; 0S5H4ZZ ; 0S5J0ZZ ; 0S5J3ZZ ; 0S5J4ZZ ; 0S5K0ZZ ; 0S5K3ZZ ; 0S5K4ZZ ; 0S5L0ZZ ; 0S5L3ZZ ; 0S5L4ZZ ; 0S5M0ZZ ; 0S5M3ZZ ;-PCS 0S5M4ZZ ; 0S5N0ZZ ; 0S5N3ZZ ; 0S5N4ZZ ; 0S5P0ZZ ; 0S5P3ZZ ; 0S5P4ZZ ; 0S5Q0ZZ ; 0S5Q3ZZ ; 0S5Q4ZZ ; 0SBH0ZZ ; 0SBH3ZZ ; 0SBH4ZZ ; 0SBJ0ZZ ; 0SBJ3ZZ ; 0SBJ4ZZ ; 0SBK0ZZ ; 0SBK3ZZ ; 0SBK4ZZ ; 0SBL0ZZ ; 0SBL3ZZ ; 0SBL4ZZ ; 0SBM0ZZ ; 0SBM3ZZ ; 0SBM4ZZ ; 0SBN0ZZ ; 0SBN3ZZ ; 0SBN4ZZ ; 0SBP0ZZ ; 0SBP3ZZ ; 0SBP4ZZ ; 0SBQ0ZZ ; 0SBQ3ZZ ; 0SBQ4ZZ ; 0MBQ0ZZ ; 0MBQ3ZZ ; 0MBQ4ZZ ; 0MBR0ZZ ; 0MBR3ZZ ; 0MBR4ZZ ; 0MTQ0ZZ ; 0MTQ4ZZ ; 0MTR0ZZ ; 0MTR4ZZ ; 0SBF0ZZ ; 0SBF3ZZ ; 0SBF4ZZ ; 0SBG0ZZ ; 0SBG3ZZ ; 0SBG4ZZ ; 0STF0ZZ ; 0STG0ZZ ; 0MBS0ZZ ; 0MBS3ZZ ; 0MBS4ZZ ; 0MBT0ZZ ; 0MBT3ZZ ; 0MBT4ZZ ; 0MTS0ZZ ; 0MTS4ZZ ; 0MTT0ZZ ; 0MTT4ZZ ; 0SBH0ZZ ; 0SBH3ZZ ; 0SBH4ZZ ; 0SBJ0ZZ ; 0SBJ3ZZ ; 0SBJ4ZZ ; 0SBK0ZZ ; 0SBK3ZZ ; 0SBK4ZZ ; 0SBL0ZZ ; 0SBL3ZZ ; 0SBL4ZZ ; 0SBM0ZZ ; 0SBM3ZZ ; 0SBM4ZZ ; 0SBN0ZZ ; 0SBN3ZZ ; 0SBN4ZZ ; 0SBP0ZZ ; 0SBP3ZZ ; 0SBP4ZZ ; 0SBQ0ZZ ; 0SBQ3ZZ ; 0SBQ4ZZ ; 0STH0ZZ ; 0STJ0ZZ ; 0STK0ZZ ; 0STL0ZZ ; 0STM0ZZ ; 0STN0ZZ ; 0STP0ZZ ; 0STQ0ZZ ; 0SGF04Z ; 0SGF05Z ; 0SGF07Z ; 0SGF0JZ ; 0SGF0KZ ; 0SGF34Z ; 0SGF35Z ; 0SGF37Z ; 0SGF3JZ ; 0SGF3KZ ; 0SGF44Z ; 0SGF45Z ; 0SGF47Z ; 0SGF4JZ ; 0SGF4KZ ; 0SGG04Z ; 0SGG05Z ; 0SGG07Z ; 0SGG0JZ ; 0SGG0KZ ; 0SGG34Z ; 0SGG35Z ; 0SGG37Z ; 0SGG3JZ ; 0SGG3KZ ; 0SGG44Z ; 0SGG47Z ; 0SGG4JZ ; 0SGG4KZ ; 0SGH04Z ; 0SGH05Z ; 0SGH07Z ; 0SGH0JZ ; 0SGH0KZ ; 0SGH34Z ; 0SGH35Z ; 0SGH37Z ; 0SGH3JZ ; 0SGH3KZ ; 0SGH44Z ; 0SGH45Z ; 0SGH47Z ; 0SGH4JZ ; 0SGH4KZ ; 0SGJ04Z ; 0SGJ05Z ; 0SGJ07Z ; 0SGJ0JZ ; 0SGJ0KZ ; 0SGJ34Z ; 0SGJ35Z ; 0SGJ37Z ; 0SGJ3JZ ; 0SGJ3KZ ; 0SGJ44Z ; 0SGJ45Z ; 0SGJ47Z ; 0SGJ4JZ ; 0SGJ4KZ ; 0SGH04Z ; 0SGH05Z ; 0SGH07Z ; 0SGH0JZ ; 0SGH0KZ ; 0SGH34Z ; 0SGH35Z ; 0SGH37Z ; 0SGH3JZ ; 0SGH3KZ ; 0SGH44Z ; 0SGH45Z ; 0SGH47Z ; 0SGH4JZ ; 0SGH4KZ ; 0SGJ04Z ; 0SGJ05Z ; 0SGJ07Z ; 0SGJ0JZ ; 0SGJ0KZ ; 0SGJ34Z ; 0SGJ35Z ; 0SGJ37Z ; 0SGJ3JZ ; 0SGJ3KZ ; 0SGJ44Z ; 0SGJ45Z ; 0SGJ47Z ; 0SGJ4JZ ; 0SGJ4KZ ; 0SGH04Z ; 0SGH05Z ; 0SGH07Z ; 0SGH0JZ ; 0SGH0KZ ; 0SGH34Z ; 0SGH35Z ; 0SGH37Z ; 0SGH3JZ ; 0SGH3KZ ; 0SGH44Z ; 0SGH45Z ; 0SGH47Z ; 0SGH4JZ ; 0SGH4KZ ; 0SGJ04Z ; 0SGJ05Z ; 0SGJ07Z ; 0SGJ0JZ ; 0SGJ0KZ ; 0SGJ34Z ; 0SGJ35Z ; 0SGJ37Z ; 0SGJ3JZ ; 0SGJ3KZ ; 0SGJ44Z ; 0SGJ45Z ; 0SGJ47Z ; 0SGJ4JZ ; 0SGJ4KZ ; 0SGK04Z ; 0SGK05Z ; 0SGK07Z ; 0SGK0JZ ; 0SGK0KZ ; 0SGK34Z ; 0SGK35Z ; 0SGK37Z ; 0SGK3JZ ; 0SGK3KZ ; 0SGK44Z ; 0SGK45Z ; 0SGK47Z ; 0SGK4JZ ; 0SGK4KZ ; 0SGL04Z ; 0SGL05Z ; 0SGL07Z ; 0SGL0JZ ; 0SGL0KZ ; 0SGL34Z ; 0SGL35Z ; 0SGL37Z ; 0SGL3JZ ; 0SGL3KZ ; 0SGL44Z ; 0SGL45Z ; 0SGL47Z ; 0SGL4JZ ; 0SGL4KZ ; 0SGM04Z ; 0SGM05Z ; 0SGM07Z ; 0SGM0JZ ; 0SGM0KZ ; 0SGM34Z ; 0SGM35Z ; 0SGM37Z ; 0SGM3JZ ; 0SGM3KZ ; 0SGM44Z ; 0SGM45Z ; 0SGM47Z ; 0SGM4JZ ; 0SGM4KZ ; 0SGN04Z ; 0SGN05Z ; 0SGN07Z ; 0SGN0JZ ; 0SGN0KZ ; 0SGN34Z ; 0SGN35Z ; 0SGN37Z ; 0SGN3JZ ; 0SGN3KZ ; 0SGN44Z ; 0SGN45Z ; 0SGN47Z ; 0SGN4JZ ; 0SGN4KZ ; 0SGH04Z ; 0SGH05Z ; 0SGH07Z ; 0SGH0JZ ; 0SGH0KZ ; 0SGH34Z ; 0SGH35Z ; 0SGH37Z ; 0SGH3JZ ; 0SGH3KZ ; 0SGH44Z ; 0SGH45Z ; 0SGH47Z ; 0SGH4JZ ; 0SGH4KZ ; 0SGJ04Z ; 0SGJ05Z ; 0SGJ07Z ; 0SGJ0JZ ; 0SGJ0KZ ; 0SGJ34Z ; 0SGJ35Z ; 0SGJ37Z ; 0SGJ3JZ ; 0SGJ3KZ ; 0SGJ44Z ; 0SGJ45Z ; 0SGJ47Z ; 0SGJ4JZ ; 0SGJ4KZ ; 0SUH0JZ ; 0SUH3JZ ; 0SUH4JZ ; 0SUJ0JZ ; 0SUJ3JZ ; 0SUJ4JZ ; 0SQF0ZZ ; 0SQF3ZZ ; 0SQF4ZZ ; 0SQFXZZ ; 0SQG0ZZ ; 0SQG3ZZ ; 0SQG4ZZ ; 0SQGXZZ ; 0SRF07Z ; 0SRF0JZ ; 0SRF0KZ ; 0SRG07Z ; 0SRG0JZ ; 0SRG0KZ ; 0MQQ0ZZ ; 0MQQ3ZZ ; 0MQQ4ZZ ; 0MQR0ZZ ; 0MQR3ZZ ; 0MQR4ZZ ; 0MQS0ZZ ; 0MQS3ZZ ; 0MQS4ZZ ; 0MQT0ZZ ; 0MQT3ZZ ; 0MQT4ZZ ; 0L8N0ZZ ; 0L8N3ZZ ; 0L8N4ZZ ; 0L8P0ZZ ; 0L8P3ZZ ; 0L8P4ZZ ; 0J8Q0ZZ ; 0J8Q3ZZ ; 0J8R0ZZ ; 0J8R3ZZ ; 0Y6P0Z0 ; 0Y6P0Z1 ; 0Y6P0Z2 ; 0Y6P0Z3 ; 0Y6Q0Z0 ; 0Y6Q0Z1 ; 0Y6Q0Z2 ; 0Y6Q0Z3 ; 0Y6R0Z0 ; 0Y6R0Z1 ; 0Y6R0Z2 ; 0Y6R0Z3 ; 0Y6S0Z0 ; 0Y6S0Z1 ; 0Y6S0Z2 ; 0Y6S0Z3 ; 0Y6T0Z0 ; 0Y6T0Z1 ; 0Y6T0Z2 ; 0Y6T0Z3 ; 0Y6U0Z0 ; 0Y6U0Z1 ; 0Y6U0Z2 ; 0Y6U0Z3 ; 0Y6V0Z0 ; 0Y6V0Z1 ; 0Y6V0Z2 ; 0Y6V0Z3 ; 0Y6W0Z0 ; 0Y6W0Z1 ; 0Y6W0Z2 ; 0Y6W0Z3 ; 0Y6X0Z0 ; 0Y6X0Z1 ; 0Y6X0Z2 ; 0Y6X0Z3 ; 0Y6Y0Z0 ; 0Y6Y0Z1 ; 0Y6Y0Z2 ; 0Y6Y0Z3 ; 0Y6M0Z4 ; 0Y6M0Z5 ; 0Y6M0Z6 ; 0Y6M0Z7 ; 0Y6M0Z8 ; 0Y6M0Z9 ; 0Y6M0ZB ; 0Y6M0ZC ; 0Y6M0ZD ; 0Y6M0ZF ; 0Y6N0Z4 ; 0Y6N0Z5 ; 0Y6N0Z6 ; 0Y6N0Z7 ; 0Y6N0Z8 ; 0Y6N0Z9 ; 0Y6N0ZB ; 0Y6N0ZC ; 0Y6N0ZD ; 0Y6N0ZF ; 0Y6M0Z0 ; 0Y6N0Z0 ; 0Y6H0Z3 ; 0Y6J0Z3 ; 0Y6H0Z1 ; 0Y6H0Z2 ; 0Y6H0Z3 ; 0Y6J0Z1 ; 0Y6J0Z2 ; 0Y6J0Z3 ; 0YMM0ZZ ; 0YMN0ZZ ; 0YMH0ZZ ; 0YMJ0ZZ ; 0YMK0ZZ ; 0YML0ZZ |
| Nerve block | At time of surgery:  CPT: 64445; 64446; 64447; 64448; 64450;  ICD-9 procedure: 04.81  ICD-10 procedure: 3E0T3BZ |
| **Physical and psychiatric comorbidities** | |
| Obesity | ICD-9 code: 278.00, 278.01;  ICD-10 code: E66.9, E66.01;  OR  Last BMI on/before index is ≥ 30 |
| Depression | ICD-9 code: 296.2x, 296.3x, 311;  ICD-10 code: F32.0-F32.5, F32.9, F33.0-F33.3, F33.4x, F33.9  - 2 outpatient occurrences (on different days) in same 12 month period or 1 inpatient occurrence. |
| Anxiety | ICD-9 code: 300.00, 300.01, 300.02, 300.23;  ICD-10 code: F41.9, F41.0, F41.1, F40.1x  - Composite of generalized anxiety disorder, panic disorder, anxiety disorder not otherwise specified, and social phobia.  - 2 outpatient occurrences (on different days) in same 12-month period or 1 inpatient occurrence on/prior to index date. |
| PTSD | ICD-9 code: 309.81;  ICD-10 code: F43.1x  - 2 outpatient occurrences (on different days) in same 12-month period or 1 inpatient occurrence on/prior to index date. |
| Obsessive compulsive disorder (OCD) | ICD-9 code: 300.3;  ICD-10 code: F42  - 2 outpatient occurrences (on different days) in same 12-month period or 1 inpatient occurrence on/prior to index date. |
| Arthritis | ICD-9 code: 710.0, 710.1, 710.2, 710.3, 710.4, 710.8, 710.9, 711.x, 713.x-717.x, 718.0x, 718.1x, 718.2x, 718.3x, 718.5x, 718.6x, 718.7x, 718.8x, 718.9x, 719.x, 720.0, V13.4;  ICD-10 code: M00.x to M02.x, M05.x, M06.x, M08.x, M12.x to M19.x, M23.x, M24.0x to M24.4x, M24.6x to M24.9, M25.x, M32.10, M33.20, M33.90, M34.0, M34.1, M34.9, M35.00, M35.01, M35.5, M35.9, M36.2, M36.3, M36.4, M43.4, M43.5x, M45.9, M79.6x, R26.2, R29.4, R29.898, Z87.39 |
| Musculoskeletal pain | ICD-9 code: 725.x, 726.0, 726.1x, 726.2, 726.3x, 726.4, 726.5, 726.6x, 726.71, 726.72, 726.90, 727.00, 727.03, 727.04, 727.05, 727.06, 727.09, 727.2, 727.3, 727.49, 727.50, 727.51, 727.6x, 727.89, 727.9, 729.0, 729.4, 729.5, 729.7x, 729.89, 729.9, 729.91, 729.92, 781.99, 830.x-848.x, 905.6, 905.7, V43.6x, V43.7. V48.3, V49.6x, V49.7x;  ICD-10 code: M35.3, M60.x to M79.x (**excluding M79.7**), R29.898, R29.91, S03.x, S13.x, S16.x, S23.x, S33.x, S39.0x, S39.9x, S43.x, S46.x, S53.x, S56.x, S63.x, S66.x, S73.x, S76.x, S83.x, S86.x, S93.x, S96.x, Z96.6x, Z97.1x, Z89.x |
| Back and Neck pain | ICD-9 code: 720.1, 720.2, 720.8x, 720.9, 721.x-722.x, 723.0-723.3, 723.5-723.7, 723.9, 724.x, 756.1x;  ICD-10 code: M43.2x, M43.6, M43.8x, M43.9, M46.0x, M46.1, M46.4x, M46.8x, M46.9x, M47.x, M48.0x, M48.1x, M48.2x, M48.3x, M48.8x, M48.9, M49.8x, M50.x to M51.x, M53.x to M54.x, M96.1, Q76.0 to Q76.3, Q76.4x, Q76.6 |
| Neuropathy | ICD-9 code: 053.13, 072.72, 337.0x, 337.1, 353.x-357.x, 377.33, 377.34, 377.41;  ICD-10 code: B02.23, B26.84, G90.0x, G99.0, G54.x to G65.x |
| Headache | ICD-9 code: 307.81, 339.x, 346.0x, 346.1x, 346.2x, 346.3x, 346.4x, 346.5x, 346.7x, 346.8x, 346.9x, 784.0;  ICD-10 code: G43.x, G44.x, R51 |
| Fibromyalgia | ICD9 - 729.1;  ICD10 – M79.7 |
| Chronic pain | ICD9 – 338.2x;  ICD10 – G89.2x |
| Smoking/nicotine dependence | ICD-9 code: V15.82, 305.1;  ICD-10 code: Z87.891, Z72.0, F17.20x, F17.21x |
| Alcohol abuse/dependence | ICD-9 code: 303.9x, 305.0x;  ICD-10 code: F10.x |
| Any drug abuse/dependence | ICD-9 code: 304.0x, 304.1x, 304.2x, 304.3x, 304.4x, 304.5x, 304.6x, 304.7x, 304.8x, 304.9x, 305.2x, 305.3x, 305.4x, 305.5x, 305.6x, 305.7x, 305.9x  ICD-10 code: F11.x, F12.x, F13.x, F14.x, F15.x, F16.x, F18.x, F19.x  - Composite of sedative, cocaine, cannabis, amphetamine, hallucinogens, ‘other,’ opioid, opioid with other SUD, other SUD excluding opioid, unspecified drug abuse/dependence. |
| **Demographics** |  |
| Age at index | Age in years |
| Race | White, African-American, other/unknown |
| Gender | Male, female |
| Census region | Midwest, Northeast, South, West, Other/unknown |
